# How good are large language models for automated data extraction from randomized trials?

**DOI:** 10.1101/2024.02.20.24303083

**Authors:** Zhuanlan Sun, Ruilin Zhang, Suhail A. Doi, Luis Furuya-Kanamori, Tianqi Yu, Lifeng Lin, Chang Xu

## Abstract

In evidence synthesis, data extraction is a crucial procedure, but it is time intensive and prone to human error. The rise of large language models (LLMs) in the field of artificial intelligence (AI) offers a solution to these problems through automation. In this case study, we evaluated the performance of two prominent LLM-based AI tools for use in automated data extraction. Randomized trials from two systematic reviews were used as part of the case study. Prompts related to each data extraction task (e.g., extract event counts of control group) were formulated separately for binary and continuous outcomes. The percentage of correct responses (*Pcorr*) was tested in 39 randomized controlled trials reporting 10 binary outcomes and 49 randomized controlled trials reporting one continuous outcome. The *Pcorr* and agreement across three runs for data extracted by two AI tools were compared with well-verified metadata. For the extraction of binary events in the treatment group across 10 outcomes, the *Pcorr* ranged from 40% to 87% and from 46% to 97% for ChatPDF and for Claude, respectively. For continuous outcomes, the *Pcorr* ranged from 33% to 39% across six tasks (Claude only). The agreement of the response between the three runs of each task was generally good, with Cohen’s kappa statistic ranging from 0.78 to 0.96 and from 0.65 to 0.82 for ChatPDF and Claude, respectively. Our results highlight the potential of ChatPDF and Claude for automated data extraction. Whilst promising, the percentage of correct responses is still unsatisfactory and therefore substantial improvements are needed for current AI tools to be adopted in research practice.

**Highlights:** *What is already known:*

- In evidence synthesis, data extraction is a crucial procedure, but it is time intensive and prone to human error, with reported data extraction error rates at meta-analyses level reaching up to 67%.
- The rise of large language models (LLMs) in the field of artificial intelligence (AI) offers a solution to these problems through automation.

*What is new:*

- In this case study, we investigated the performance of two AI tools for data extraction and confirmed that AI tools can reach the same or better performance than humans in terms of data extraction from randomized trials for binary outcomes.
- However, AI tools performed poorly at extracting data from continuous outcomes.

*Potential impact for Research Synthesis Methods readers outside the authors’ field:*

- Our study suggests LLMs have great potential in assisting data extraction in evidence syntheses through (semi-)automation. Further efforts are needed to improve accuracy, especially for continuous outcomes data.

## 1 Introduction

The recent expansion of ChatGPT and other AI assistance tools have generated considerable attention in the healthcare domain ^1,2^. ChatGPT is a large language model (LLM) developed by OpenAI and relies on a large corpus of text data to pre-train on billions of parameters using a transformer model. It generates coherence and human-like text responses and considers the contextual information of preceding words based on given prompts ^3^. Researchers have explored the potential of using ChatGPT for various tasks, including improving the efficiency and effectiveness of clinical management ^4–6^, and supporting healthcare services ^7–9^, hereby benefiting patients and medical practitioners ^10–12^.

With the wide utilization of ChatGPT, more well-known LLM-based AI tools such as ChatPDF (based on ChatGPT) ^13^ and Claude (Version 2, another AI tool powered by Anthropic’s LLM) ^14^ specialize in extracting information from uploaded PDF files and offer a promising opportunity for evidence synthesis. A critical process in evidence synthesis is data extraction, which serves as the cornerstone for generating credible evidence. However, data extraction is time-intensive ^15^ and susceptive to human errors ^16^. In reproducibility studies, data extraction errors were found to occur frequently, with an error rate of 17.0% at the study level and 66.8% at the meta-analysis level ^17^. Such errors call into question the credibility of evidence syntheses and their usefulness for health care practice, leading to consequences such as wrong conclusions leading to poorer decisions. AI technology has the potential to reduce the manual workload required for data extraction in evidence syntheses and reduce human errors ^18^. In this case study, we evaluated the performance of two stand-out LLM-powered AI tools, ChatPDF and Claude, for automatic data extraction from randomized controlled trials (RCTs). We focussed on accuracy and agreement by extracting information from a predefined set of questions provided as prompts for ChatPDF and Claude.

## 2 Methods

### 2.1 Selection of sample reviews

We started our research by selecting appropriate ‘cases’. We chose the data source (systematic reviews of RCTs) based on two considerations: 1) the original systematic reviews should be free of data extraction error after our further checking; 2) the original systematic reviews should be published in reputational journals. Two systematic reviews of randomized trials published in *BMJ* and *Sleep Medicine Reviews* were selected, with one reporting binary outcomes ^19^ and another reporting continuous outcomes ^20^. The first systematic review investigated the risk of neuropsychiatric adverse events associated with varenicline based on 39 randomized controlled trials (RCTs) and 10 adverse outcomes (e.g., depression, fatigue and anxiety) ^19^. The second systematic review investigated the effects of non-pharmacological sleep interventions on depressive symptoms based on 49 RCTs and one efficacy outcome (depression) ^20^. The metadata of these two systematic reviews were extracted by the review authors and verified by our research team ^17,21^.

### 2.2 Data extraction tasks

For binary outcomes, the 2 by 2 table data (i.e., event counts, group size in each arm) of each trial for a specific outcome was of interest for extraction. For continuous outcomes, the 3 by 2 table data (i.e., mean, standard deviation, group size in each arm) of each trial for a specific outcome was of interest for extraction. Each cell of the above table data refers to a specific task; therefore, there were 4 tasks for binary outcomes and 6 tasks for continuous outcomes within each trial (Table S1).

### 2.3 AI tools for data extraction

There were two steps for AI tools for the data extraction tasks. First, appropriate prompts (questions or statements to interact with AI) for each task were formulated; this was done by referring to a standardized and regulated set ^22^. Second, these prompts were inputted into ChatPDF and Claude to process the data extraction tasks. Considering that the initially designed question/statements for automated data extraction may not be the optimal one, we employed ChatGPT to modify the prompts by leveraging the aforementioned type of prompts, such as “Please design the best prompt for me based on this prompt: …”.

Given that the responses may vary for duplicated attempts even with the same question, we conducted three separate runs for each question on each RCT article. All the responses of ChatPDF and Claude were collected in mid-September 2023. Each prompt was conducted in a new session to alleviate memory retention bias.

### 2.4 Outcomes

The primary outcome was the percentage of correct extractions (*Pcorr*) for each task, which was measured by number of correctly extracted RCTs over the total number of RCTs to be extracted. For example, in a task for extracting the death events of the intervention group in 10 RCTs, when correct extractions of the number of deaths were achieved in 5 RCTs, then the *Pcorr* for the outcome was 50%. The secondary outcome was the percentage of incorrect extractions (*Picorr*) and the rate of percentage that failed to identify the information (*Pfail*). *Picorr* and *Pfail* for each task were measured by calculating the ratio of incorrectly and unsuccessfully extracted RCTs to the total number of RCTs to be extracted, respectively. The sum of *Pcorr*, *Picorr,* and *Pfail* for each task is equal to 1. The test-retest reliability of each question based on three separate runs, as mentioned above, was also presented as a measure of the robustness of each question and was categorized as follows: slight, fair, moderate, substantial, and almost perfect agreement based on the following value ranges: [0.00, 0.20], [0.21, 0.40], [0.41, 0.60], [0.61, 0.80], and [0.81, 1.00], respectively [19].

### 2.5 Statistical analysis

The *Pcorr*, *Picorr*, and *Pfail* proportions of each task were measured for each of the 11 outcomes. Since group sizes were mostly trial dependent, the tasks for group sizes extraction (i.e., treatment and control) were tested directly based on the 39 trials regardless of which of the 10 binary outcomes was involved. For the tasks of event counts, the three proportions (*Pcorr*, *Picorr*, and *Pfail*) were computed for each of the 10 outcomes and then synthesized across the 10 outcomes for a weighted mean via the inverse variance heterogeneity method (IVhet) with Freeman-turkey double arcsine transformation of the proportions ^23,24^. The test-retest reliability was measured via the *Kappa* statistic, which accounts for the potential impact of chance on agreement ^25^. The statistical analysis was conducted using the “DescTools” package in R and MetaXL 5.3 (EpiGear International Pty Ltd.) ^26^.

## 3 Results

Figure 1 illustrates the use of LLM-based AI tools for data extraction. Table 1 presents the details of the 11 outcomes. The original and modified prompts are presented in Tables S2 and S3, and examples of prompt responses for both ChatPDF and Claude are presented in Table S4. Since there were no obvious differences in the *Pcorr* of the tasks under different types of prompts (i.e., original question-based prompt, original statement-based prompt, modified question-based prompt, and modified statement-based prompt), here we only report the results of the original question-based prompt. The results for the other three prompts are displayed in Figures S1-S3.

**Figure 1.**
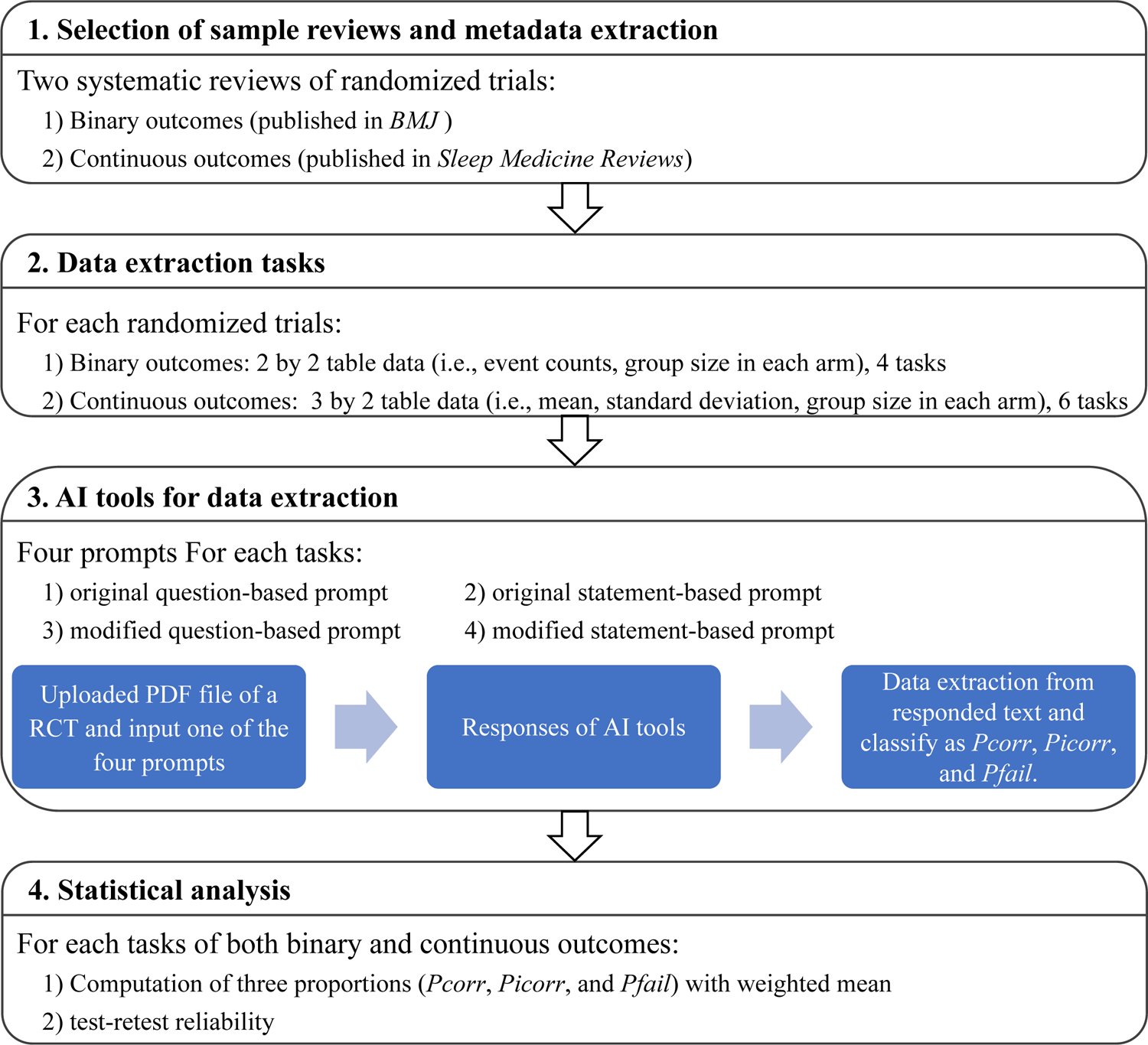
Flowchart of using LLM-based AI tools for data extraction.

**Table 1.**
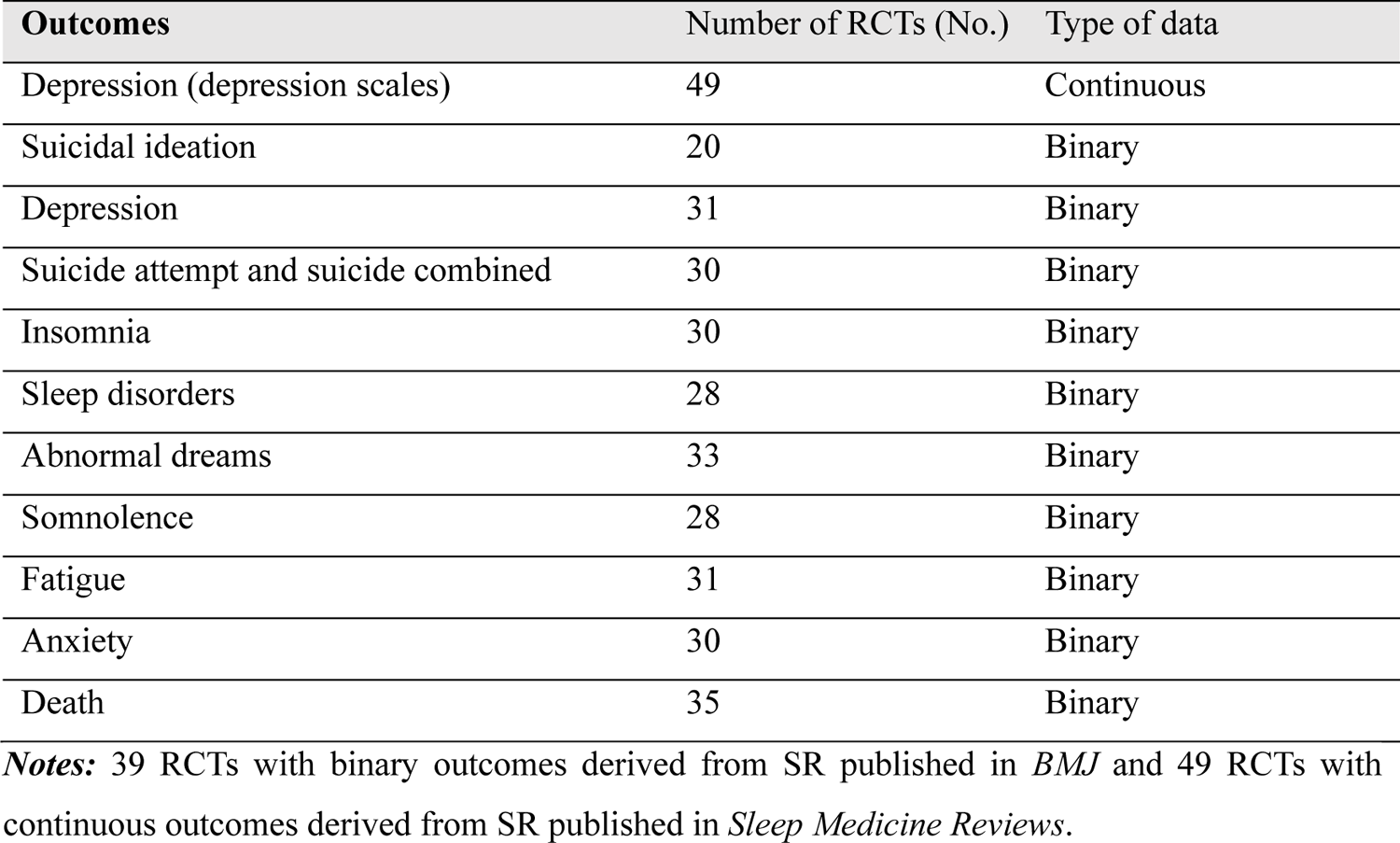
General characteristics of data collected from RCTs.

### 3.1 Binary outcomes

#### 3.1.1 Group size information

The *Pcorr* for extracting the group size information in the treatment group (Task 1) and control group (Task 2) across 39 trials based on ChatPDF was 54% (95%CI: 38%, 69%) and 59% (95%CI: 43%, 74%) respectively (Figure 2). The *Pcorr* when using Claude for the two tasks across trials was 72% (95%CI: 57%, 85%) and 77% (95%CI: 62%, 89%). For the *Picorr* and *Pfail*, they were lower in ChatPDF and Claude for all tasks; see Figure S4.

**Figure 2.**
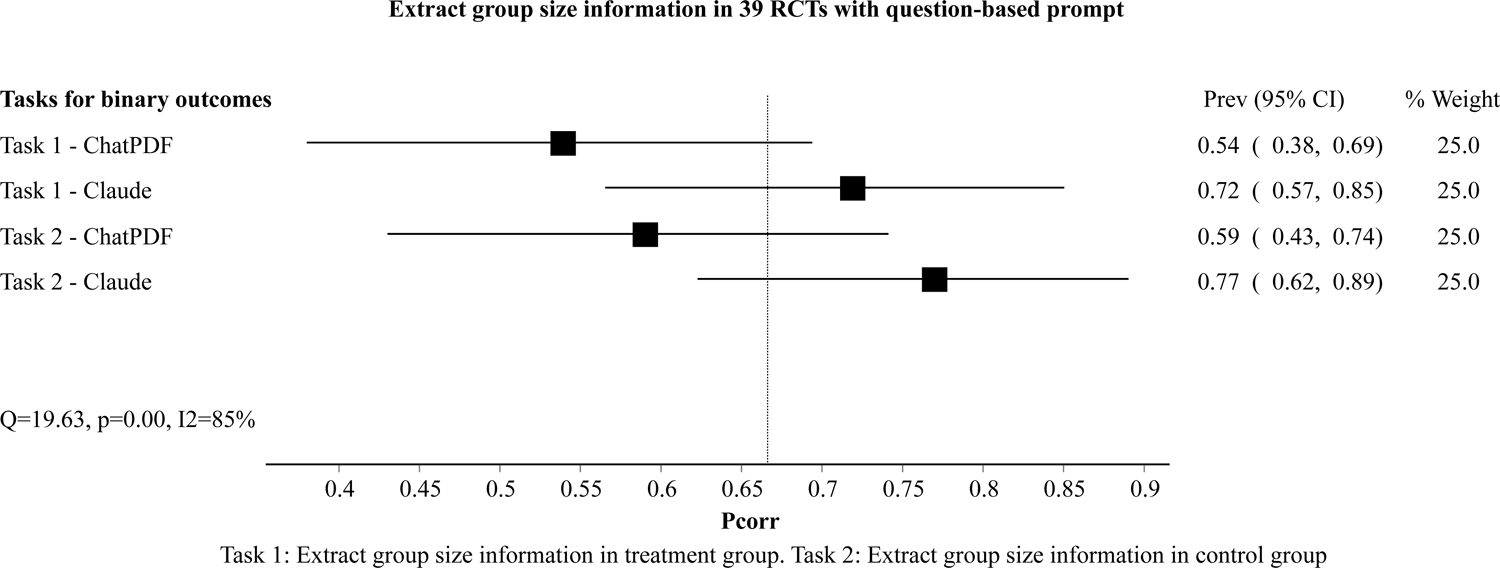
Correct responses (*Pcorr*) of AI tools for group size data extraction from RCTs with binary outcomes.

#### 3.1.2 Event counts

Figure 3 presents the performance on task 3 (extracting the event count in the treatment group). The *Pcorr* of ChatPDF for extracting the event count ranged from 40% to 87% across 10 outcomes, with a weighted mean *Pcorr* of 64% (95%CI: 57%, 68%, I^2^ = 84%). The *Pcorr* when using Claude for extracting the event count ranged from 46% to 97%, with a weighted mean *Pcorr* of 70% (95%CI: 65%, 76%, I^2^ = 69%).

**Figure 3.**
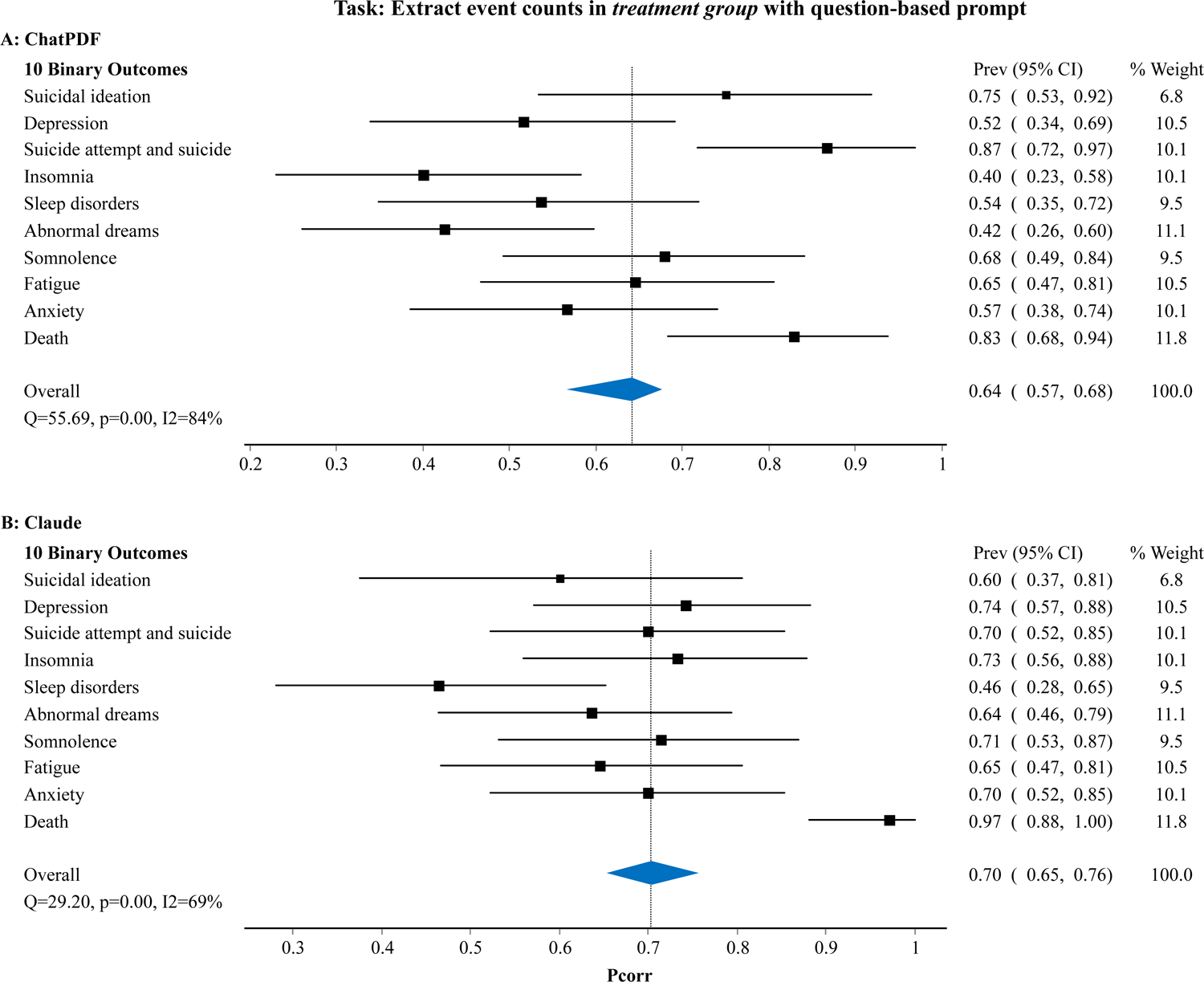
Correct responses (*Pcorr*) of AI tools for extracting event counts in treatment group.

Figure 4 presents the performance on task 4 (extracting the event count in the control group). The *Pcorr* of ChatPDF for extracting the event count ranged from 33% to 86% across 10 outcomes, with a weighted mean of 64% (95%CI: 56%, 67%, I^2^ = 86%). The *Pcorr* of using Claude for extracting the event count ranged from 50% to 97%, with a weighted mean of 75% (95%CI: 70%, 80%, I^2^ = 71%). Again, *Picorr* and *Pfail* were lower in ChatPDF and Claude across all tasks; see Figures S5 and S6.

**Figure 4.**
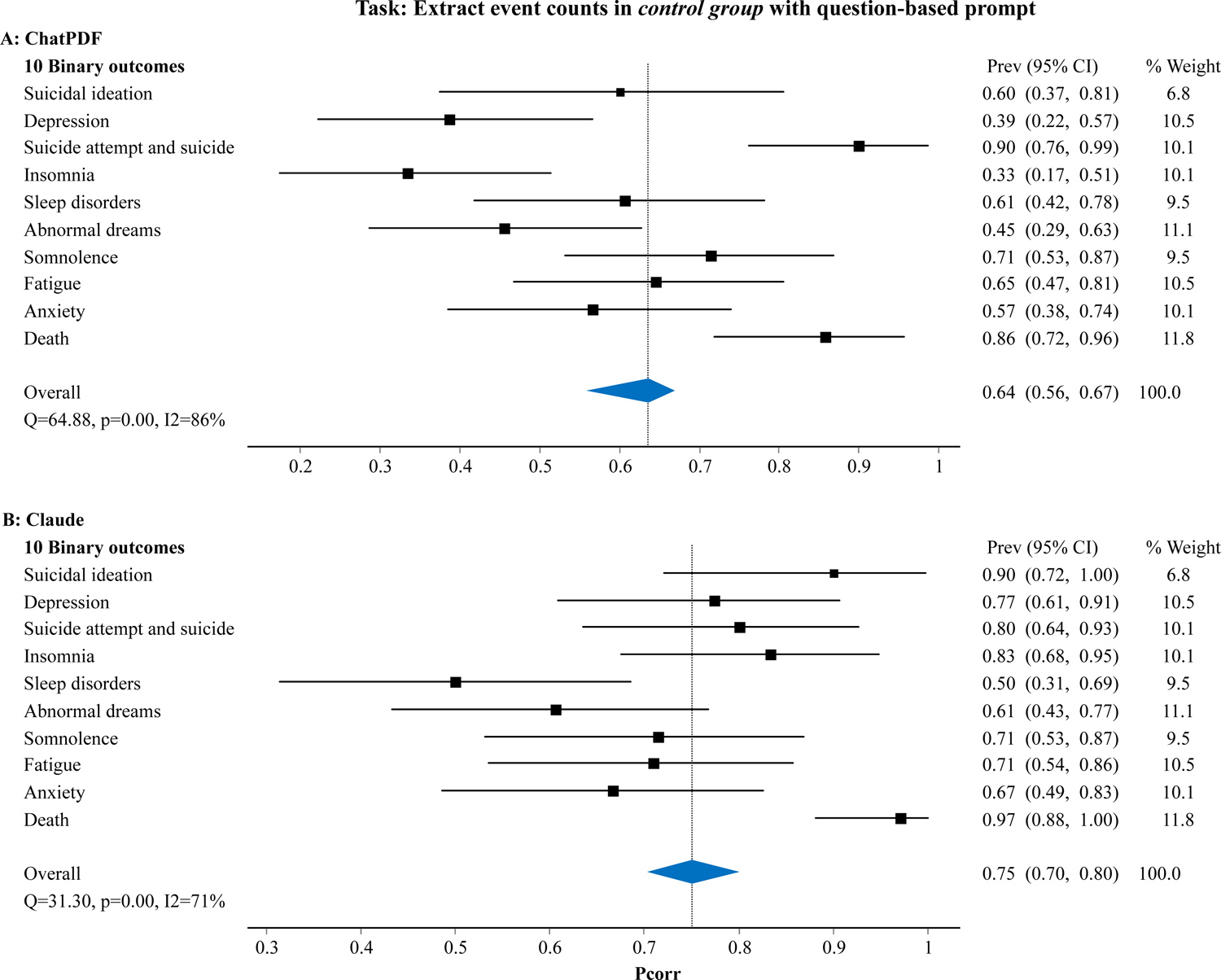
Correct responses (*Pcorr*) of AI tools for extracting event counts in control group.

### 3.2 Continuous outcomes

There was only one continuous outcome, and thus the *Pcorr* were reported for the six tasks and no synthesis was needed. Using Claude, the accuracy for extracting the mean value in treatment group (Task 1), standard deviation in treatment group (Task 2), group size in treatment group (Task 3), mean value in control group (Task 4), standard deviation in control group (Task 5), group size in control group (Task 6) were 39%, 39%, 33%, 33%, 37%, and 37% (see Figure 5). The *Pcorr* was not tested in ChatPDF since it failed to extract data from complicated tables. The *Picorr* and *Pfail* with Claude were presented in Figure S7.

**Figure 5.**
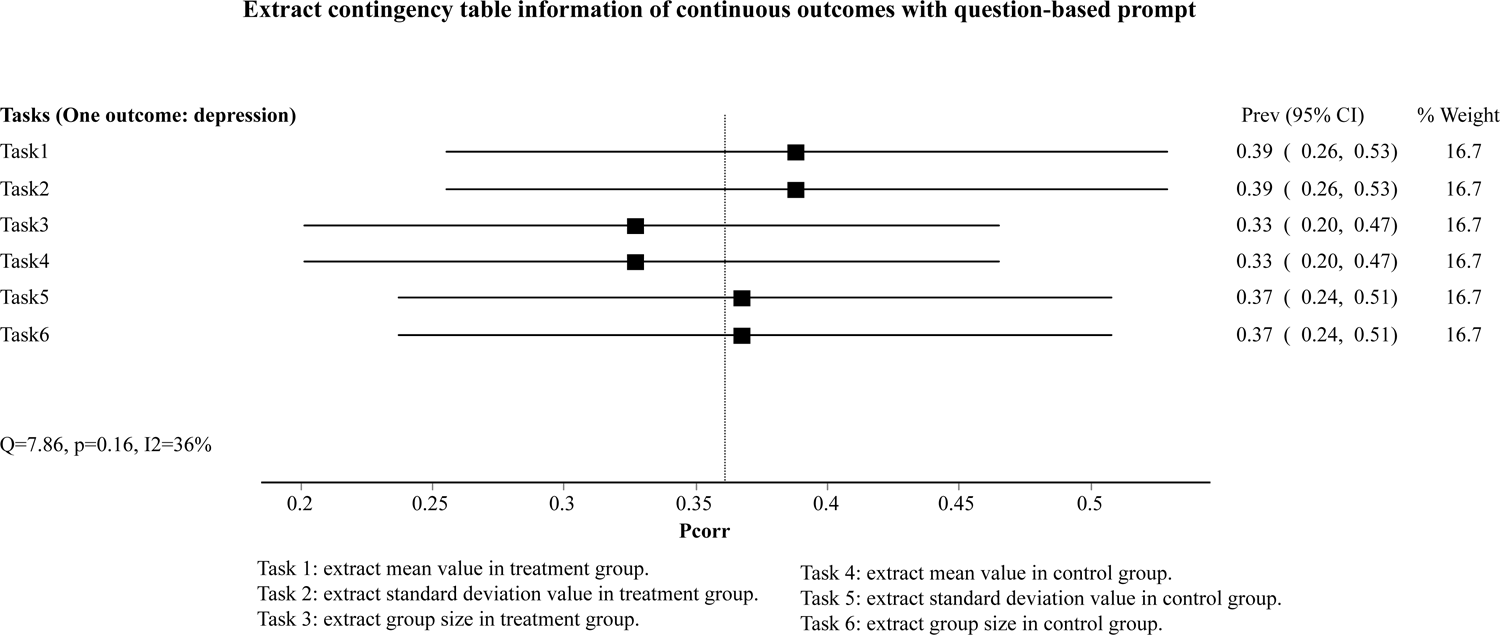
Correct responses (*Pcorr*) of AI tool (Claude only) for data extraction from RCTs with continuous outcomes.

### 3.3 Test-retest reliability analysis of automation data extraction

Cohen’s kappa coefficients are presented in Table 2. For binary outcomes, the *Kappa* coefficients of the responses of the three runs of each question-based prompt ranged from 0.94 to 0.96 for ChatPDF and 0.78 to 0.92 for Claude, demonstrating a nearly perfect level of concordance. For continuous outcomes, the *Kappa* values of the responses of the three runs of each question-based prompt ranged from 0.65 to 0.82 for Claude, which is substantial but not perfect.

**Table 2.** *Kappa* for testing the test-retest reliability of each prompt.

| Prompt IDs | Cohen's kappa coefficient | Lower 95% CI | Upper 95% CI |
| --- | --- | --- | --- |
| **Binary outcomes** |  |  |  |
| *ChatPDF* |  |  |  |
| P1 (Original) | 0.94 | 0.92 | 0.96 |
| P2 (Original) | 0.93 | 0.91 | 0.96 |
| P1 (Modified) | 0.95 | 0.92 | 0.97 |
| P2 (Modified) | 0.96 | 0.93 | 0.98 |
| *Claude* |  |  |  |
| P1 (Original) | 0.78 | 0.76 | 0.79 |
| P2 (Original) | 0.80 | 0.79 | 0.82 |
| P1 (Modified) | 0.90 | 0.88 | 0.92 |
| P2 (Modified) | 0.92 | 0.90 | 0.94 |
| **Continuous outcomes** |  |  |  |
| *Claude* |  |  |  |
| P3 (Original) | 0.78 | 0.76 | 0.79 |
| P4 (Original) | 0.82 | 0.80 | 0.83 |
| P3 (Modified) | 0.65 | 0.64 | 0.66 |
| P4 (Modified) | 0.72 | 0.71 | 0.73 |

**Notes:** *Kappa* coefficients are categorized as slight, fair, moderate, substantial, and almost perfect agreement based on the value ranges of [0.00, 0.20], [0.21, 0.40], [0.41, 0.60], [0.61, 0.80], and [0.81, 1.00], respectively.

## 4 Discussion

In our investigation of the performance of two AI tools (ChatPDF and Claude), we found that their correct responses in the automatic data extraction of binary outcomes are generally good, reaching as high as 75%. This is promising when compared to humans’ performance. In our recent randomized trial, the accuracy of data extraction for safety outcomes by humans was about 65% in terms of single extraction and 75% in terms of double extraction for the same tasks ^27^. These results serve to underscore the potential utility of AI tools for automating data extraction in evidence synthesis of binary outcomes.

However, for continuous data, the performance of the AI tool was poor, with only 39% correct responses. This is mainly due to the high failure rate (over 40%) to identify the information (see Figure S4). In randomized trials, the reporting of the mean value and standard deviation of a continuous outcome was generally more complex, with fewer locator words or phrases to posit the target number, and often involved missing values (i.e., report median value and quartile range instead of mean and standard deviation) ^29^. These were the reasons that resulted in the high failure rate. Thus, in evidence synthesis of continuous data, current AI technology may not be accurate enough to assist with the data extraction process.

Three reasons may contribute to the unsatisfactory accuracy of automation data extraction by using ChatPDF and Claude. First, They are primarily designed to summarize an article’s key findings. Though the latest version of ChatPDF can identify pages containing specific information, ChatPDF and Claude can sometimes misidentify information from the text and tables of an article, and even fail to identify data contained in complicated tables, resulting in inaccurate data extraction and even no response. We observe this limitation of using both ChatPDF and Claude for automated data extraction from RCTs with continuous outcomes, where most of the data are presented in tables. Second, ChatPDF relies on ChatGPT 3.5 for now, and Claude is sensitive to changes in prompts ^30^, which results in the generation of different responses even when presented with the same prompt multiple times (see the varied *Pcorr* in Figures S8-S10) ^31^. Also, reformulating the prompts may result in different responses. This has been shown in the test-retest reliability results (Table 2). Third, ChatGPT and Claude leverage Internet-scale corpus and curated knowledge base as training data; neither was specifically trained on biomedical corpora.

Although tools for data extraction in systematic reviews have been extensively investigated, the usability of these automation tools depends on awareness of their existence and familiarity with their functionalities ^28^. To overcome barriers to the utilization of these tools, user-friendly design as well as comprehensive training and support that enhances the usability of the tools are indispensable. Importantly, a higher accuracy rate of automated data extraction is the priority consideration for the use of the tools in evidence synthesis.

Some limitations of our study should be noted. We used a relatively small sample size, involving the analysis of data extraction accuracy from only 88 RCTs included in two SR articles published in a specific journal. The representativeness might be limited. Thus, exploring more relevant papers becomes a worthwhile consideration for future research. Moreover, ChatPDF (which currently relies on ChatGPT 3.5) shares similarities with ChatGPT. This may impose certain limitations, such as the potential for inaccuracies in generating information. In addition, we observed variability in ChatPDF and Claude responses to the same prompt across different iterations, despite deliberately designing three prompts and their optimized versions using ChatGPT. Further research is essential to systematically investigate the impact of prompt variations on data extraction accuracy. This exploration should encompass not only additional AI tools, such as Bard and Bing ^32^, but also a more extensive pipeline considering the incorporation of tools like langchain to optimize the overall process. A further limitation of this study is that no prompts were tested on extracting other data that can be useful such as medians, ranges, IQRs, and *p*-values (particularly in situations where the other data are not reported and reviewers might need such data to estimate other data for meta-analysis). Encouragement is extended for further research endeavors aimed at addressing this research gap and enhancing the depth of exploration in this research direction.

## 5. Conclusions

Based on the evidence of our case study, we found that current LLM-powered AI tools could achieve good correct responses for data extraction tasks for binary outcomes, while for continuous outcomes, the AI tool performed poorly. Our study suggests the great potential of utilizing large language models to assist evidence synthesis in automating data extraction for binary outcomes. To ensure the accuracy of automated data extraction, it is still prudent to rely solely on AI tools. It is recommended to combine the use of AI tools with human extraction to enhance the efficiency and accuracy of automation data extraction. Further efforts are still needed to improve the accuracy of the data extraction, especially for continuous outcomes data.

## Contributions

ZLS conceived the study, performed and interpreted the analyses, and wrote the first draft manuscript; RLZ performed and interpreted the analyses; CX conceived the study, reviewed and interpreted the analyses, and made major contributions to the writing of the manuscript, and approved the final version of the manuscript. SD, LFK, TQY, and LFL provide methodological guidance and revised the draft. All authors read and approved the final version of the manuscript.

## Data Availability

All data produced in the present work are contained in the manuscript

## Acknowledgments

Not applicable.

## Data sharing Statement

The dataset used in the analysis are shared at OSF (https://osf.io/6w8cz/).

